# Germline pathogenic variants in 786 neuroblastoma patients

**DOI:** 10.1101/2023.01.23.23284864

**Authors:** Jung Kim, Zalman Vaksman, Laura E. Egolf, Rebecca Kaufman, J. Perry Evans, Karina L. Conkrite, Arnavaz Danesh, Gonzalo Lopez, Michael P. Randall, Maiah H. Dent, Lance M. Farra, Neil Menghani, Malwina Dymek, Heena Desai, Ryan Hausler, Penn Medicine BioBank, Regeneron Genetics Center, Cancer Genomics Research Laboratory, Jaime Guidry Auvil, Daniela S. Gerhard, Hakon Hakonarson, Kara N. Maxwell, Kristina A. Cole, Trevor J. Pugh, Kristopher R. Bosse, Javed Khan, Jun S. Wei, John M. Maris, Douglas R. Stewart, Sharon J. Diskin

## Abstract

**Importance:** Neuroblastoma accounts for 12% of childhood cancer deaths. The genetic contribution of rare pathogenic germline variation in patients without a family history remains unclear.

**Objective:** To define the prevalence, spectrum, and clinical significance of pathogenic germline variation in cancer predisposition genes (CPGs) in neuroblastoma patients.

**Design, Setting and Participants:** Germline DNA sequencing was performed on the peripheral blood from 786 neuroblastoma patients unselected for family history. Rare variants mapping to CPGs were evaluated for pathogenicity and the percentage of cases harboring pathogenic (P) or likely pathogenic (LP) variants was quantified. The frequency of CPG P-LP variants in neuroblastoma cases was compared to two distinct cancer-free control cohorts to assess enrichment. Matched tumor DNA sequencing was evaluated for “second hits” at CPGs and germline DNA array data from 5,585 neuroblastoma cases and 23,505 cancer-free control children was analyzed to identify rare germline copy number variants (CNVs) affecting genes with an excess burden of P-LP variants in neuroblastoma. Neuroblastoma patients with germline P-LP variants were compared to those without P-LP variants to test for association with clinical characteristics, tumor features, and patient survival.

**Main Outcomes and Measures:** Rare variant prevalence, pathogenicity, enrichment, and association with clinical characteristics, tumor features, and patient survival.

**Results:** We observed 116 P-LP variants in CPGs involving 13.9% (109/786) of patients, representing a significant excess burden of P-LP variants compared to controls (9.1%; P = 5.14 × 10^−5^, Odds Ratio: 1.60, 95% confidence interval: 1.27-2.00). *BARD1* harbored the most significant burden of P-LP variants compared to controls (1.0% vs. 0.03%; P = 8.18 × 10^−7^; Odds Ratio: 32.30, 95% confidence interval: 6.44-310.35). Rare germline CNVs disrupting *BARD1* were also identified in neuroblastoma patients (0.05%) but absent in controls (P = 7.08 × 10^−3^; Odds Ratio: 29.47, 95% confidence interval: 1.52 – 570.70). Overall, P-LP variants in DNA repair genes in this study were enriched in cases compared to controls (8.1% vs. 5.7%; P = 0.01; Odds Ratio: 1.45, 95% confidence interval: 1.08-1.92). Neuroblastoma patients harboring a germline P-LP variant had a worse overall survival when compared to patients without P-LP variants (P = 8.6 × 10^−3^), and this remained significant in a multivariate Cox proportional-hazards model (P = 0.01).

**Conclusions and Relevance:** Neuroblastoma patients harboring germline P-LP variants in CPGs have worse overall survival and *BARD1* is an important predisposition gene affected by both common and rare pathogenic variation. Germline sequencing should be performed for all neuroblastoma patients at diagnosis to inform genetic counseling and support future longitudinal and mechanistic studies. Patients with a germline P-LP variant should be closely monitored, regardless of risk group assignment.

**Key Points:** 

**Question:** What is the prevalence and clinical significance of germline pathogenic variants in cancer predisposition genes (CPGs) in neuroblastoma patients?

**Findings:** Among 786 neuroblastoma patients with germline DNA sequencing, 13.9% harbored a pathogenic (P) or likely pathogenic (LP) variant in a CPG. The number of patients with germline P-LP variants in *BARD1* and other DNA repair genes was significantly greater than observed in two cancer-free control cohorts. The presence of a germline P-LP variant was independently predictive of worse overall survival.

**Meaning:** Germline sequencing should be performed for all neuroblastoma patients at diagnosis to inform genetic counseling and frequency of clinical follow-up. Centralization of these data will facilitate longitudinal and mechanistic studies needed to identify specific actionable events and improve patient outcomes.

## Introduction

Neuroblastoma is an embryonal malignancy of early childhood that arises from developing postganglionic sympathetic neurons and accounts for 12% of all childhood cancer-related deaths^1^. Patients are classified into low-, intermediate- and high-risk based on a series of clinical and tumor biological features and this risk group is used for treatment stratification purposes^1^. Despite aggressive multi-modal therapy, nearly 50% of high-risk neuroblastoma patients diagnosed over 18 months of age eventually succumb to their disease. A subset of these tumors harbor somatic *MYCN* amplification and/or an activating somatic *ALK* mutation or gene amplification^1^. However, sequencing studies of neuroblastoma tumors have revealed a low overall somatic mutation rate and few recurrently mutated genes^2,3^. The young median age at diagnosis and standardized incidence ratio of siblings of children with neuroblastoma of ∼9.7^4^ are consistent with an underlying genetic etiology.

The genetic basis of neuroblastoma predisposition has come into focus over the last decade. Familial neuroblastoma, which accounts for 1-2% of cases, arises primarily from pathogenic germline variants in *ALK*^5^, with rarer neurocristopathy syndrome cases explained by germline pathogenic variants in *PHOX2B*^6,7^.

However, the vast majority of neuroblastomas appear to arise sporadically, without a family history. Genome-wide association studies (GWAS) have identified common variants associated with sporadic neuroblastoma at over a dozen loci. These genetic associations have implicated multiple candidate genes including *CASC15, NBAT1, BARD1, LMO1, DUSP12, DDX4, IL31RA, HSD17B12, HACE1, LIN28B, TP53, RSRC1, MLF1, CPZ, MMP20, KIF15*, and *NBPF23*^8-16^. Several susceptibility genes identified by GWAS not only influence disease initiation, but also drive tumor aggressiveness and/or maintenance of the malignant phenotype^10,12,14,17-20^. A rare 16p11.2 microdeletion syndrome has also been associated with neuroblastoma^21^. Finally, recent sequencing efforts have reported rare pathogenic germline variants in multiple cancer predisposition genes^2,22-32^; however, the prevalence and clinical significance of these and other rare variants in neuroblastoma remains unclear and requires evaluation in larger patient cohorts with detailed phenotypic data.

Here, we analyzed germline whole genome sequencing (WGS), whole exome sequencing (WES) and targeted capture sequencing (CAP) data from 786 children diagnosed with neuroblastoma and profiled through the Therapeutically Applicable Research to Generate Effective Treatments (TARGET) initiative. Our aims were to 1) determine the prevalence, spectrum, and pathogenicity of rare germline variants in known cancer predisposition genes (CPGs), 2) test for enrichment of rare variants in children with neuroblastoma compared to cancer-free control populations, and 3) evaluate clinical features and outcomes in neuroblastoma patients with and without germline pathogenic (P) or likely pathogenic (LP) variants in CPGs to identify translational opportunities.

## Materials and Methods

Detailed methods are provided in **Supplemental Methods**. Briefly, the study cohort consisted of 786 neuroblastoma patients accrued through the Children’s Oncology Group (COG) ANBL00B1 biology study, unselected for family history (**Table 1, eTable 1**). Patients are assigned a Universal Subject Identifier (USI) by the COG and are not identifiable. Germline DNA and matched diagnostic tumor DNA and RNA were sequenced through the TARGET initiative. The original set of tumor-normal pairs were sequenced with Complete Genomics WGS (n=134) and/or Illumina WES (n=222), as previously described^2,33^. A total of 59 samples were sequenced by WGS and WES provided internal validation. We have previously reported a small number of germline variants based on the WES cohort^2^; however, an in-depth study of pathogenic germline variation in these children was not performed at that time. Germline DNA from an independent neuroblastoma cohort (n=489) was sequenced using Illumina custom capture (CAP) panels, including a germline panel newly designed for this study (n=166 genes; **eTable 2**). Cancer-free control data were obtained from the Penn Medicine BioBank (PMBB; n=6,295)^34,35^ and gnomAD v2.1 without cancer (n=15,708). Ancestry for neuroblastoma and PMBB participants was inferred through principal component analysis using matched germline DNA array data. Germline variants were called using GATK best practices (WES and CAP data) or the Complete Genomics pipeline (v2) with custom filtering^36^ (WGS data), then annotated with SnpEff^37^ (v4.3t) and ANNOVAR^38^. For all cohorts (neuroblastoma, PMBB and gnomAD), rare germline variants (<0.1% across each population in public control databases) in 166 cancer predisposition genes were then assessed for pathogenicity with a clinically-focused pipeline incorporating ClinVar evidence and a modified implementation of InterVar (**eFigure 1**). A subset of germline variants were validated through Sanger sequencing. Cases harboring a germline P-LP variant in a cancer predisposition gene were further assessed using matched tumor sequencing data when available. Fisher’s exact test was used to compare the enrichment of P-LP variants in neuroblastoma cases to controls and across clinical and biological subsets of neuroblastoma. P-LP variant enrichment was also compared at gene and pathway levels, using a Bonferroni correction for multiple testing. Kaplan-Meier analyses of event-free and overall survival were performed to compare outcomes of patients with and without germline P-LP variants. A multivariate Cox proportional-hazards regression model was used to assess if the presence of a CPG P-LP variant was independently predictive of survival.

**Table 1.**
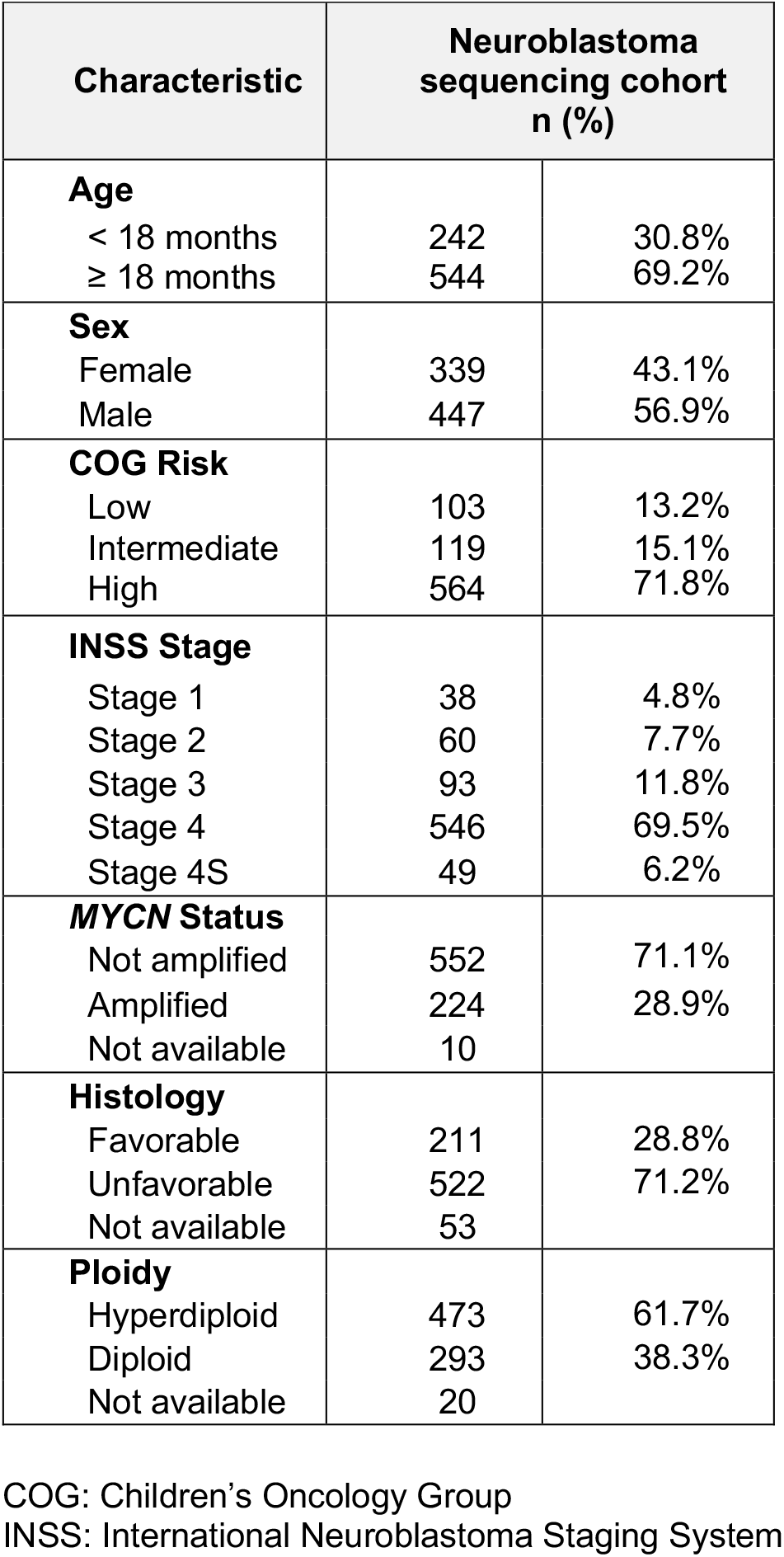
Clinical and tumor biological characteristics for 786 neuroblastoma cases.

| Characteristic | Neuroblastoma sequencing cohort<br>n (%) |  |
| --- | --- | --- |
| **Age** |  |  |
| < 18 months | 242 | 30.8% |
| ≥ 18 months | 544 | 69.2% |
| **Sex** |  |  |
| Female | 339 | 43.1% |
| Male | 447 | 56.9% |
| **COG Risk** |  |  |
| Low | 103 | 13.2% |
| Intermediate | 119 | 15.1% |
| High | 564 | 71.8% |
| **INSS Stage** |  |  |
| Stage 1 | 38 | 4.8% |
| Stage 2 | 60 | 7.7% |
| Stage 3 | 93 | 11.8% |
| Stage 4 | 546 | 69.5% |
| Stage 4S | 49 | 6.2% |
| **MYCN Status** |  |  |
| Not amplified | 552 | 71.1% |
| Amplified | 224 | 28.9% |
| Not available | 10 |  |
| **Histology** |  |  |
| Favorable | 211 | 28.8% |
| Unfavorable | 522 | 71.2% |
| Not available | 53 |  |
| **Ploidy** |  |  |
| Hyperdiploid | 473 | 61.7% |
| Diploid | 293 | 38.3% |
| Not available | 20 |  |

COG: Children's Oncology Group
INSS: International Neuroblastoma Staging System

## Results

### Neuroblastoma patient characteristics

A total of 786 neuroblastoma patients were included in the study (**Table 1, eTable 2**). Overall, 564 cases (71.8%) were classified as high-risk based on the COG risk stratification system, and 546 of these patients had stage 4 disease according to the International Neuroblastoma Staging System (INSS) criteria. Cases profiled by WGS or WES were intentionally enriched for high-risk disease, consistent with the overall goals of the TARGET initiative. In contrast, cases that underwent targeted capture (CAP) sequencing were representative of the general neuroblastoma risk group profile. A total of 769 patients had available matched germline DNA array data and were evaluated for ancestry by principal component analysis (PCA). As expected, the majority of cases (66.8%) were inferred to be of European ancestry (**eFigure 2**).

### Frequency of P-LP variants in known cancer predisposition genes

We observed 116 P-LP variants involving 54 of the 166 cancer predisposition genes studied (**Figure 1, eTable 3**). Of these variants, 73 were classified as P-LP based on ClinVar evidence and 43 variants were assigned P-LP based on our revised InterVar assessment. Overall, P-LP variants were detected in 109/786 (13.9%) of cases. Classic familial neuroblastoma germline variants were observed in 0.4% (3/786) of cases. These included two patients with *ALK* (p.R1275Q) activating variants and a single patient with a *PHOX2B* splice variant (NM_003924:exon3:c.430-2A>G). An additional *ALK* variant (p.I1250T) was predicted to be LP; however, this variant was not found to be activating in a previous study^39^. Six cases harbored more than one P-LP variant in a known cancer predisposition gene (**eTable 4**). A total of 27 genes harbored P-LP variants in two or more cases, with variants in the *BARD1* gene being most frequent (8/786 cases or 1.0% overall; **eFigure 3, eTable 3**). Select variants were validated by Sanger sequencing (**eTable 5**). Notably, of the nine neuroblastoma genes identified by GWAS and included in this study, only *BARD1* and *TP53* harbored rare P-LP variants.

**Figure 1.**
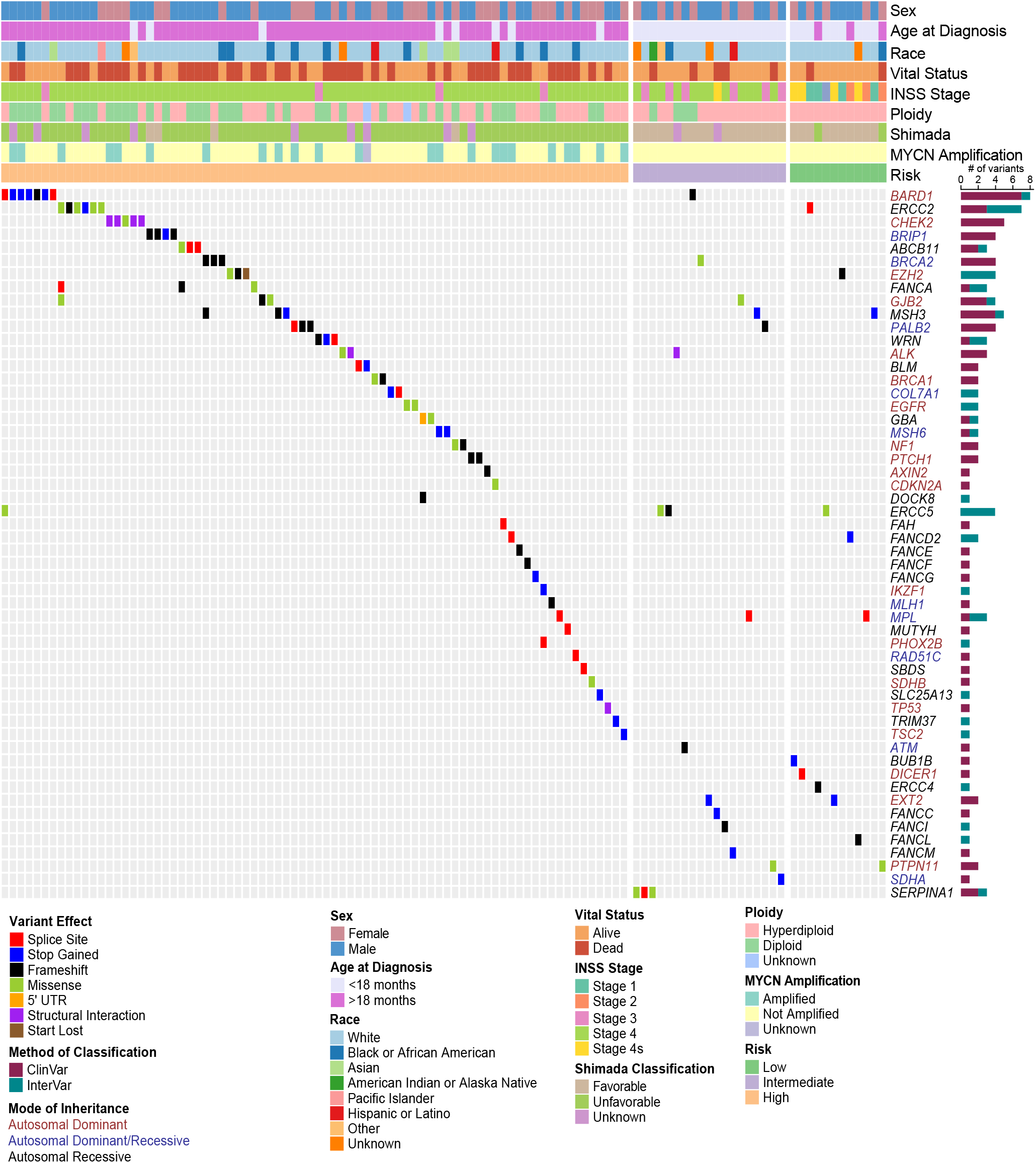
Germline P-LP variants in cancer predisposition genes across 786 neuroblastoma cases. Oncoprint of known cancer predisposition genes (CPGs) harboring rare germline variants classified as pathogenic (P) or likely pathogenic (LP). Samples without P-LP variants in these genes are not shown. Samples are ordered by COG risk group and annotated with patient clinical and tumor biologic features. Genes are color-coded according to mode of inheritance, when known. Bar chart to the right indicates the number of variants detected for each gene and whether pathogenicity was determined based on ClinVar or our modified InterVar automated assessment. All variants were manually reviewed for quality and evidence of pathogenicity.

### Matched tumor DNA sequencing reveals P-LP variants are retained and second hits are rare

We examined somatic alterations affecting genes with germline P-LP variants using matched published neuroblastoma tumor WGS and WES variant calls^33^ and our analysis of available CAP sequencing from TARGET. Nearly all germline P-LP variants were detected in matched tumor DNA, when available (95%, 73/77 variants; **eTable 3**). For variants not detected in the matched tumor (n=4), germline variant allele fraction (VAF) ranged from 0.25 to 0.31. No second hit SNVs or indels were observed in the tumor DNA of patients harboring a germline P-LP variant in a CPG. We detected one focal somatic deletion within *EZH2* observed in the tumor from PASEGA, a patient who also harbored a pathogenic germline *EZH2* variant (p.T536fs). Phase could not be determined, but this somatic event was confirmed by Sanger sequencing of the tumor DNA (**eFigure 4**).

### Germline P-LP variants in cancer predisposition genes are enriched in neuroblastoma

To assess whether children diagnosed with neuroblastoma harbor an excess of rare pathogenic germline variation in the 166 CPGs considered in this study, we compared the P-LP burden in neuroblastoma cases to two independent control cohorts (**Figure 2A, top panel**). First, we applied our full analytic pipeline (alignment, variant calling, quality control, and pathogenicity assessment) to subjects sequenced through the Penn Medicine Bio Bank (PMBB) and without a history of cancer or benign tumors (n=6,295, **eFigure 5**). Germline P-LP variants were significantly enriched in neuroblastoma patients compared to PMBB (P_PMBB_ = 5.14 × 10^−5^; Odds Ratio: 1.60; 95% confidence interval: 1.27-2.00). To assess reproducibility of this result, identical filtering and pathogenicity assessment was applied to rare variants in gnomAD excluding cancer samples (n=15,708 individuals). This confirmed the excess burden of P-LP variants in neuroblastoma (P_gnomAD_ = 1.82 × 10^−3^; Odds Ratio: 1.41; 95% confidence interval: 1.34-1.74).

**Figure 2.**
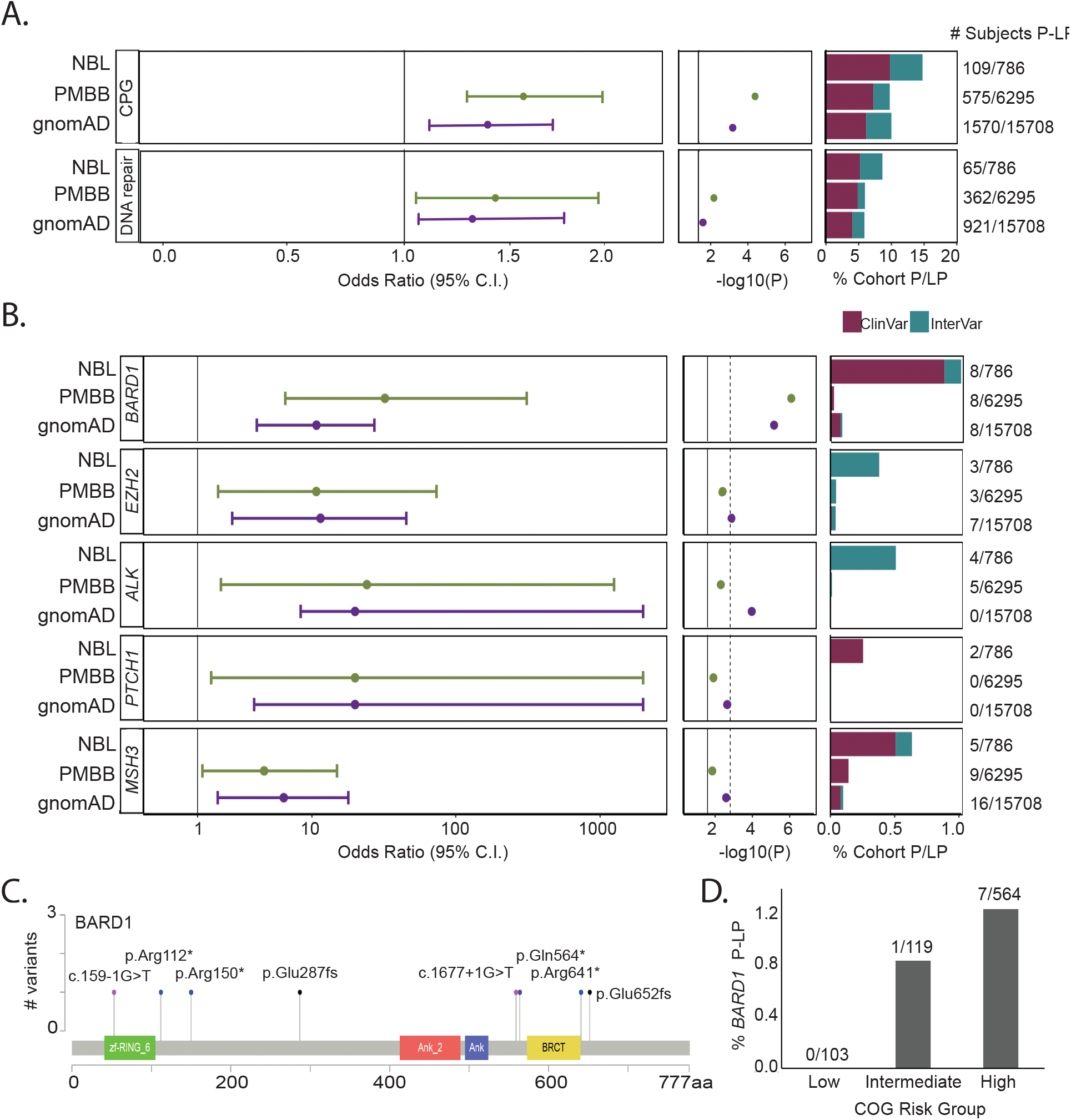
Neuroblastoma cases harbor an excess burden of rare P-LP variants in cancer predisposition genes. **A**. Overall excess burden of P-LP variants (SNVs and indels) in neuroblastoma cases vs. PMBB and gnomAD v2.1 controls is shown for cancer predisposition genes and the subset of genes studied involved in DNA repair. **B**. Gene-based rare variant burden test results comparing the number of neuroblastoma subjects with P-LP variants to those detected in gnomAD v2.1 and PMBB. **C**. Lollipop figure depicting eight germline P-LP variants in BARD1. **D**. Rare P-LP variants in BARD1 are observed predominantly in patients diagnosed with high-risk neuroblastoma.

### Germline P-LP variants in BARD1 are enriched in neuroblastoma patients

Next, we performed gene-based rare variant burden testing comparing neuroblastoma cases to PMBB and gnomAD control cohorts (**eTable 6, eTable 7**). Five genes (*BARD1, EZH2, ALK, PTCH1* and *MSH3*) exhibited significant enrichment (P < 0.05) and odds ratio lower 95% confidence interval > 1.0 in both control cohort comparisons (**Figure 2B**). Pathogenic variants in *BARD1* and *EZH2* were validated by Sanger sequencing in neuroblastoma cases when DNA was available (**eFigure 3** and **eFigure 4**). *ALK* and *BARD1* remained significant after Bonferroni adjustment for multiple testing in at least one control comparison. *ALK* is the main major familial neuroblastoma predisposition gene^5^. Only one LP variant in *ALK* was detected in PMBB (P_PMBB_ = 5.00 × 10^−3^; Odds Ratio: 24.09, 95% confidence interval: 1.93-1255.78). This variant (p.A1168T) was classified LP based on InterVar and was not reported in ClinVar. No P-LP *ALK* variants were observed in gnomAD v2.1 whole genome controls (P_gnomAD_ = 1.08 × 10^−4^; Odds Ratio: 140.3, 95% confidence interval: 7.24-2719.0).

*BARD1* is the only gene that passed a Bonferroni adjustment in both control comparisons (**Figure 2B**). Common variation at the *BARD1* locus is known to be associated with high-risk neuroblastoma from our prior GWAS^9^. We and others have also reported germline *BARD1* rare variants in neuroblastoma patients^2,40,41^.

However, to date, the number of cases analyzed has been limited. Here, we observed rare P-LP variants in *BARD1* in 8/786 (1.0%) of neuroblastoma cases, all predicted to be loss-of-function (**Figure 2C** and **eTable 8**). Moreover, all but one variant was observed in the high-risk subset (**Figure 2D**). Only 2/6,295 (0.03%) of control subjects in PMBB harbored a P-LP germline variant in *BARD1* (P_PMBB_ = 8.18 × 10^−7^; Odds Ratio: 32.30, 95% confidence interval: 6.44-310.35). Similarly, only 15/15,708 (0.09%) control subjects in gnomAD harbored a P-LP germline variant in *BARD1* (P_gnomAD_ = 6.64 × 10^−6^; Odds Ratio: 10.75, 95% confidence interval: 3.93-27.13). Enrichment of P-LP variants in *BARD1* remained significant when we restricted the analysis to cases and controls of European ancestry, considering both PMBB (P_PMBB_= 2.70 × 10^−5^; Odds Ratio: 21.36, 95% confidence interval: 4.55-131.80) and gnomAD (P_gnomAD_ = 1.55 × 10^−4^; Odds Ratio: 10.13, 95% confidence interval: 3.75-28.74.86), suggesting this result is not likely due to population stratification.

### Germline P-LP variants in DNA repair genes are enriched in neuroblastoma patients

*BARD1* is known to bind *BRCA1* and influence DNA repair^42^. We hypothesized that neuroblastoma patients harbor an excess burden of P-LP variants in DNA repair pathway genes overall. To explore this hypothesis, we interrogated genes in our 166-gene panel that intersected published DNA repair genes.^43^ A total of 48 DNA repair genes were assayed in the full study cohort and included in the analysis (**eTable 2**). We observed 68 P-LP variants in 27 distinct DNA repair genes, affecting 64/786 (8.1%) of neuroblastoma patients (**Figure 2A, bottom panel**). In contrast, only 362/6,295 (5.8%) of PMBB controls harbored a P-LP variant (P_PMBB_ = 0.011; Odds Ratio: 1.45, 95% confidence interval: 1.08-1.92). Similarly, only 959/15,708 (6.1%) of gnomAD controls harbored a P-LP variant in a DNA repair gene (P_gnomAD_ = 0.028; Odds Ratio: 1.36, 95% confidence interval: 1.03-1.77).

### Germline copy number variants (CNVs) disrupting BARD1 are enriched in neuroblastoma

Given the enrichment of *BARD1* rare P-LP SNVs and indels in this study and the previous association of *BARD1* common variants with neuroblastoma through GWAS, we next sought to determine if rare germline CNVs at *BARD1* also associate with neuroblastoma. We analyzed CNVs in germline DNA array data from 5,585 neuroblastoma cases and 23,505 cancer-free control children genotyped in our neuroblastoma GWAS efforts^21^. We detected three focal germline deletions fully or partially encompassing *BARD1* in neuroblastoma (0.05%; **Figure 3A-E, eTable 9**). No CNVs affecting *BARD1* were observed in 23,505 chip-matched GWAS controls, and no protein coding deletions affecting *BARD1* were observed in 10,847 individuals in the gnomAD v2.1 structural variant dataset^44^ (**Figure 3A**). The rare germline deletions at *BARD1* were significantly associated with neuroblastoma (P = 7.08 × 10^−3^; Odds Ratio: 29.47, 95% confidence interval: 1.52 – 570.70; **Figure 3F**) and were detected only in high-risk neuroblastoma patients (**Figure 3G**). Collectively, these data suggest that *BARD1* alterations are an important genetic determinant of neuroblastoma, including common germline variants and rare germline SNVs, indels, and structural variants.

**Figure 3.**
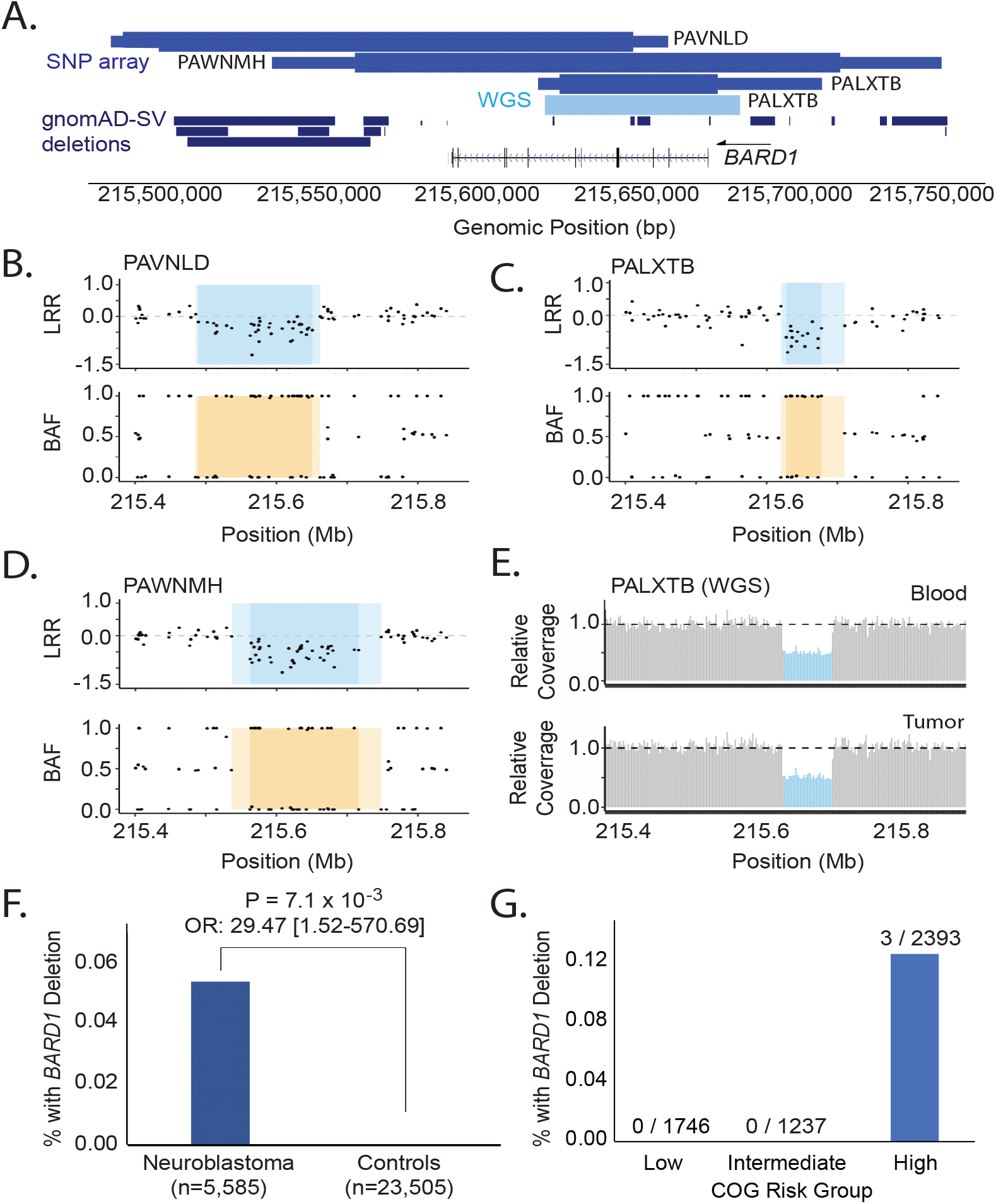
Rare germline CNVs disrupting *BARD1* in neuroblastoma. **A**. *BARD1* deletions were identified in 3 out of 5,585 neuroblastoma patients through copy number analysis of a large germline DNA array dataset (medium blue, top track). No deletions were observed in 23,505 chip-matched controls. The thick and thin bars represent minimum and maximum deletion coordinates, respectively. One deletion was validated and fine-mapped by WGS (light blue, middle track). No *BARD1* protein coding deletions were observed in 10,847 individuals in the gnomAD v2.1 structural variant dataset (dark blue, bottom track). **B-D**. The three array-based CNV calls are shown in log R ratio (LRR) and B allele frequency (BAF) plots. Darker shading indicates the minimum deleted region, whereas lighter shading indicates the maximum region. **E**. WGS validation for patient PALXTB is shown as relative sequencing coverage for matched blood and tumor samples. **F**. Rare *BARD1* deletion CNVs are enriched in neuroblastoma compared to cancer-free controls. **G**. Deletions disrupting *BARD1* were observed exclusively in patients diagnosed with high-risk subset of neuroblastoma.

### Neuroblastoma patients harboring germline P-LP variants in CPGs have worse overall survival

Finally, we investigated whether rare P-LP variants (SNVs and indels) in CPGs were associated with specific clinical and tumor biological characteristics and patient survival. A nominally significant enrichment of P-LP variants was observed in patients with tumors harboring loss of heterozygosity (LOH) of chromosome 11q (P = 0.012); however no association with age at diagnosis, stage, *MYCN* amplification status, COG risk group, or other characteristics was detected (**eTable 10**). We repeated this analysis using the CAP data only; however, the results were similar (**eTable 11**). We next evaluated overall survival probability based on the presence or absence of germline P-LP variants in CPGs. We observed that subjects with a germline P-LP variant have worse overall survival compared to subjects without a germline P-LP variant in a CPG (log-rank test P = 8.6 × 10^−3^; **Figure 4A**). Furthermore, if restricted to only low- and intermediate-risk subjects, overall survival remains worse for patients with a germline P-LP variant (log-rank test: P= 1.3 × 10^−4^; **Figure 4B**). A similar trend was observed when restricted to high-risk only, though this did not reach statistical significance (log-rank test P = 0.1049; **Figure 4C**). Finally, a multivariate Cox proportional-hazards regression model revealed the presence of a germline P-LP variant was independently predictive of overall survival when considering age and diagnosis, INSS stage, *MYCN* amplification status, and COG risk group (**Figure 4D**; p=0.017; hazard ratio= 1.44, 95% confidence interval: 1.07-1.96; **eTable 12**). Taken together, these data demonstrate that the presence of a germline P-LP variant in a CPG is associated with worse survival, independent of risk group.

**Figure 4.**
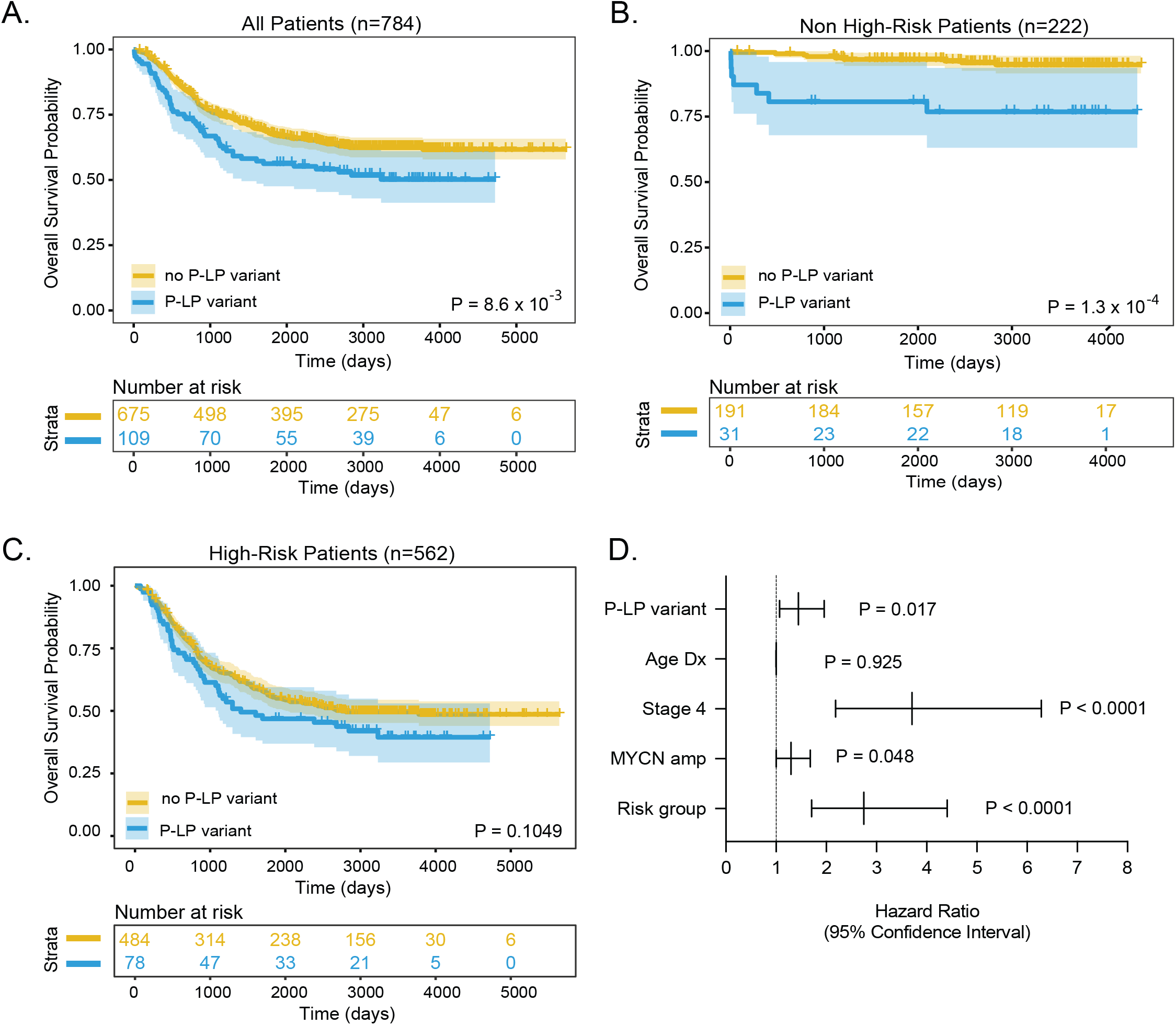
Patient carriers of germline P-LP variants in CPGs have worse outcome. Kaplan-Meier plots of overall survival probability in neuroblastoma patients with and without P/LP variants cancer predisposition genes. **A**. All patients **B**. Restricted to low- and intermediate risk (non-high-risk) groups. **C**. Restricted to high-risk group. Statistical significance in A-C assessed by log-rank test (P < 0.05). **D**. Forest plot of hazard ratios from Cox proportional-hazards model.

## Discussion

Neuroblastoma is a cancer of the developing sympathetic nervous system with an established genetic basis. Patients who present with a family history of the disease most commonly harbor rare pathogenic variants in *ALK*^5^ or *PHOX2B*^6,7^. In contrast, GWAS studies have identified common variation associated with sporadic neuroblastoma implicating over a dozen susceptibility genes^45^, including *BARD1*^9^. Germline sequencing studies have reported rare pathogenic variation in CPGs, including *APC, AXIN2, BARD1, BRCA1, BRCA2, CHEK2, LZTR1, PALB2, PINK1, SDHB, SMARCA4*, and *TP53*^*46*^. Neuroblastoma has also been reported in several childhood-onset tumor-predisposition syndromes^46^. However, a study of germline pathogenic variants in a clinically annotated cohort of patients large enough to investigate excess burden of pathogenic variation and clinical features associated with such variation has not been previously reported. Thus, in this study, we sought to define the prevalence, spectrum, and clinical significance of rare pathogenic germline variants in CPGs in neuroblastoma.

To accomplish these goals, we analyzed germline DNA sequencing from 786 neuroblastoma patients with detailed clinical covariate and outcomes data. Using a conservative, clinically-focused pipeline to classify pathogenicity of rare variants in cancer predisposition genes, we observed P-LP germline variants in a substantial portion of cases studied (13.9%). This percentage is slightly higher, but overall in-line with recent pan-childhood cancer germline studies^22,23,28,31,32,40^. Two genes (*ALK* and *BARD1*) showed enrichment of P-LP variants in neuroblastoma cases compared to independent cancer-free control cohorts after adjusting for multiple testing. Neuroblastoma patients carrying a germline P-LP variant had worse overall survival compared to those without P-LP variants, independent of age at diagnosis, INSS stage, *MYCN* amplification, and COG risk group.

The greatest number of pathogenic germline variants were observed in *BARD1, BRCA2, ERCC2, CHEK2*, and *MSH3*. Notably, *BARD1, BRCA2, CHEK2* and *MSH3* variants were primarily classified as P-LP based on ClinVar annotation, suggesting that these variants have previously been observed in patients in a clinical lab. All five genes are involved in DNA repair, and indeed we observed an overall enrichment of P-LP variants in DNA repair genes considered here. While this finding requires validation using a full repertoire of DNA repair genes, the result suggests that neuroblastoma is another cancer initiated by germline defects in DNA repair. The current study was large enough to demonstrate a significant enrichment of rare *BARD1* P-LP variants (SNVs and CNVs) in neuroblastoma, adding to the common variants in *BARD1* identified by GWAS and previously implicated in disease pathogenesis. In addition to *BARD1*, we observed 1.7% (13/786) of children in our cohort with a P-LP variant in *BRCA1, BRCA2* or a mismatch repair gene, which is similar to that observed (∼1.2%) in a large (n=3,975) meta-analysis of childhood cancer studies^47^. Notably, two of these subjects harbored multiple P-LP variants in these genes, including one patient with a P-LP variant in both *BRCA1* and *BRCA2* another patient with P-LP variants in *BRCA2* and *MSH3*.

In a companion study (co-submitted), multiple lines of evidence demonstrated the impact of *BARD1* germline P-LP variants identified here on DNA repair processes in neuroblastoma^48^. Briefly, a subset of the rare *BARD1* variants identified in the current study were introduced as monoallelic “knock-ins” in neuroblastoma cell models via CRISPR/Cas9 genome editing. These heterozygous variants induced *BARD1* haploinsufficiency leading to DNA repair deficiency and increased DNA damage. Taken together, these data further implicate *BARD1* and DNA repair as important driving factors in neuroblastoma tumorigenesis.

Evidence for bi-allelic inactivation and/or loss of heterozygosity (LOH) in patients with P-LP germline variants was observed in only one tumor and involved *EZH2*. There are several possible explanations for the low rate of bi-allelic inactivation in neuroblastoma. First, haploinsufficiency may be sufficient to tumorigenesis, as seen in our companion *BARD1* functional studies (co-submitted)^48^. Alternatively, other inactivation mechanisms may be present but not detected by our approach (*e*.*g*., epigenetic and non-coding alterations). Finally, there have been limited number of subjects with germline P-LP variants and matched tumor data evaluated to date.

Functional studies, such as those presented for *BARD1*^*48*^, and large cohort analyses incorporating the full spectrum of potential inactivation mechanisms are needed to resolve these important questions.

Given the prevalence and relevance of germline pathogenic variants to survival in neuroblastoma, we propose that all newly diagnosed children should undergo paired germline-tumor sequencing and efforts should be made to centralize these data. This is important for multiple reasons: 1) from a prognostic standpoint, identification of a P-LP variant predicts worse overall survival, independent of clinical risk stratification, 2) identification of P-LP variants in some genes (e.g. *BARD1*) may suggest eligibility for specific therapies, especially at time of relapse, 3) cascade testing of adult family members may guide gene-specific surveillance and therapies (e.g., in *BARD1, BRCA1, BRCA2, CHEK2*, Lynch, *PALB2* and *TP53*), 4) cascade testing of children for select genes (e.g., *TP53, PTPN11, DICER1*) may guide surveillance and therapy and, 5) identification of pathogenic variation in genes associated with specific syndromes (e.g., *PTCH1*/Gorlin, *EZH2*/Weaver) may guide prognosis and clinical management. While the field is adopting paired germline-tumor DNA sequencing at the time of diagnosis for neuroblastoma and other pediatric cancer patients, there remains confusion on how to act on findings. Centralization of these data, through the Childhood Cancer Data Initiative (CCDI) or similar efforts, will facilitate longitudinal studies of patient survival and provide a resource for prioritizing functional and mechanistic studies to identify specific actionable insights to improve outcomes.

There are some limitations to this study. Patients studied were enrolled in the North American neuroblastoma biology study, ANBL00B1. These cases are predominantly of European ancestry, and may not be representative of other geographic locations and ancestries. Second, while we attempted to control for this in our burden testing, the use of different sequencing methods (WGS, WES, CAP) may affect capture and variant detection at some genetic loci. Third, non-coding variants, epigenetic alterations and an exhaustive set of structural variants were not analyzed here due to the different sequencing methods used. Finally, due to lack of parental DNA, we cannot say whether the P-LP identified in this study are inherited or acquired *de novo*.

Additional large studies from diverse populations, and including parental DNA, are needed to replicate and extend our results.

In conclusion, this study of 786 neuroblastoma patients found that 13.9% harbor rare germline pathogenic variants in one or more CPGs. Rare pathogenic variants (SNVs and CNVs) in *BARD1* and other DNA repair genes were significantly enriched in neuroblastoma compared to cancer-free controls. The presence of one or more germline P-LP variants in a CPG was independently associated with worse overall survival. These data may be used to inform decision-making regarding genetic testing and potential therapeutic options for children diagnosed with neuroblastoma.

## Data Availability

Source data analyzed in this study are available through the database of Genotypes and Phenotypes (dbGaP) by requesting access to the following accession numbers:
Sequencing data: phs000467
DNA Array data: phs000124
All additional data and results in the present study are available upon reasonable request to the authors

https://www.ncbi.nlm.nih.gov/projects/gap/cgi-bin/study.cgi?study_id=phs000124.v3.p1

https://www.ncbi.nlm.nih.gov/projects/gap/cgi-bin/study.cgi?study_id=phs000467.v21.p8

## Acknowledgments

This work was supported by a supplement to the Children’s Oncology Group Chair’s grant CA098543 and with federal funds from the National Cancer Institute, National Institutes of Health, under Contract No.

HHSN261200800001E to S.J.D and Complete Genomics. This work was also supported by National Institutes of Health grants R01CA204974 (S.J.D.), R01CA237562 (S.J.D.), R03CA230366 (S.J.D.), R35CA220500 (J.M.M), K08CA230223 (K.R.B.), a Howard Hughes Medical Institute Medical Fellows grant (M.P.R.), Alex’s Lemonade Stand Foundation (K.R.B.), the EVAN Foundation (K.R.B.), and the Intramural Research Program of the Division of Cancer Epidemiology and Genetics and Center for Cancer Research of the National Cancer Institute, Bethesda, MD. K.R.B. is a Damon Runyon Physician-Scientist supported (in part) by the Damon Runyon Cancer Research Foundation (PST-07-16). This work used resources of the NIH High Performance Computing Biowulf cluster and the Children’s Hospital of Philadelphia High Performance Compute cluster.

