## Supplementary Information for "Germline pathogenic variants in 786 neuroblastoma patients"

#### I. SUPPLEMENTARY METHODS

##### *Neuroblastoma patient samples*

A total of 786 neuroblastoma patients accrued through the North American-based Children's Oncology Group (COG) ANBL00B1 biology study were included in the study (**Table 1**, **eTable 1**). Patients were unselected for family history of neuroblastoma. Genomic DNA was extracted from peripheral blood lymphocyte samples and obtained through the COG nucleic acids bank housed at the Children's Hospital of Philadelphia (CHOP). Matched diagnostic tumor DNA and RNA was also obtained from the same biobank. The patient cohort was intentionally enriched for high-risk disease and poor outcome through the Therapeutically Applicable Research to Generate Effective Treatments (TARGET) initiative. We have previously reported a small number of germline variants based on exome sequencing in a subset (n=222) of these patients[1]; however, an in-depth study of pathogenic germline variation in these children was not performed at that time.

##### *Penn Med Biobank (PMBB)*

The Penn Medicine BioBank (PMBB) is a precision medicine cohort with genomic profiling of participants who consented for biospecimen collection and linkage of their biospecimen to their electronic health record (EHR) data.[2] Starting in 2004, participants were recruited into PMBB at the time of medical appointments in the University of Pennsylvania Health System. All individuals recruited were patients of clinical practice sites of the University of Pennsylvania Health System. Appropriate consent was obtained from each participant regarding storage of biological specimens, genetic sequencing, access to all available EHR data, and permission to recontact for future studies. The study was approved by the Institutional Review Board of the University of Pennsylvania.

Phenotyping and identification of PMBB participants who were cancer-free and also had no history of a benign tumor was performed as described previously.[3] Briefly, participants with cancer were identified from the EHR using International Classification of Diseases (ICD)-9 or ICD-10 billing codes. These included both prevalent and incident cases. We then defined a cancer- and tumor-free control cohort as individuals with no ICD-9/10 codes for invasive cancer, benign, in situ, or secondary neoplasms (n=6,295).

In this study, we included 6,295 individuals who had undergone whole-exome sequencing (WES) and germline genome-wide DNA array-based genotyping. For each individual, DNA was extracted from stored buffy coats and then exome sequences were generated by the Regeneron Genetics Center (Tarrytown, NY) and obtained for study. For purposes of this study, all WES data were reprocessed as described above for neuroblastoma data. Germline genome-wide DNA array-based genotyping using the Infinium Global Screening Array (GSA) chip (Illumina) was utilized to infer ancestry of the 6,295 individuals in the cancer- and tumor-free control cohort. Genotyping was performed at the Children's Hospital of Philadelphia or Regeneron Genetics Center as previously described.[3]

***Ancestry inference***

Paired germline single nucleotide polymorphism (SNP) array data were utilized to infer ancestry of neuroblastoma and Penn Medicine Biobank (PMBB) control subjects. Genotypes from neuroblastoma cases were intersected with data form the International HapMap Project (HapMap v3, draft release 2). The variants were pruned using a window size of 50 variants, step size of five variants, and pairwise $r^2$ threshold of 0.2. A principal component analysis (PCA) was performed using PLINK 1.9 and ancestry inferred as previously described[4]. SNP array data from PMBB samples were processed in an identical manner. Neuroblastoma SNP array data are available through the Database of Genotypes and Phenotypes (dbGaP), accession phs000124.v3.p1.

***Genomic sequencing***

Genomic DNA was sequenced using a combination of Complete Genomics whole genome sequencing (WGS), Illumina whole exome sequencing (WES), and Illumina targeted capture sequencing (CAP). Complete Genomics WGS (n=134) and Illumina WES (n=222) of matched tumor-normal DNA pairs was generated through the TARGET initiative (https://ocg.cancer.gov/programs/target/data-matrix), as previously described[1,5]. A total of 76 tumor-normal pairs were profiled by both WES and WGS, allowing for internal validation (**eTable 1**). An independent set of tumor-normal pairs (n=499) were sequenced by Illumina-based targeted capture sequencing within the TARGET project. Since this TARGET capture panel was designed to include genes known to be somatically mutated in childhood cancer, we constructed a new custom capture panel to include known cancer predisposition genes, neuroblastoma syndrome-related genes, and candidate susceptibility genes identified by GWAS (n=166 genes; **eTable 2**). We performed CAP germline-only sequencing of 489 of the 499 neuroblastoma cases with DNA available. For this CAP sequencing, 200 ng genomic DNA from peripheral blood lymphocyte samples was purified using Agencourt AMPure XP Reagent (Beckman Coulter Inc, Brea, CA, USA) and a next-generation sequencing library was prepared with the KAPA HyperPlus Kit (KAPA Biosystems, Wilmington, MA) using Bioo Scientific NEXTflex™ DNA Barcoded Adapters (Bioo Scientific, Austin, TX, USA) according to KAPA-provided protocol. Libraries were barcoded, amplified, pooled, and captured using NimbleGen's SeqCap EZ Choice Library with custom-designed 523-gene panel (Roche NimbleGen, Inc., Madison, WI, USA). Captured pooled libraries were amplified, cleaned, quantified, and sequenced on a HiSeq2000 using 2x125 bp paired-end sequencing protocols (Illumina, San Diego, CA, USA). Average depth of coverage for targeted capture regions across all subjects was 198x. Concordance with TARGET CAP germline sequencing was assessed for genes that overlapped in the custom captures.

##### *Germline variant calling and annotation*

Variants for Complete Genomics WGS data were produced from Complete Genomics pipeline (v2) in hg19 and single nucleotide variants (SNVs) were filtered as previously described[6]. A decision tree classifier was designed and implemented to retain high-confidence small insertion and deletion (indel) calls from this pipeline. Illumina WES (neuroblastoma and PMBB controls) and CAP (neuroblastoma) data were aligned to hg19 using Burrows Wheeler Aligner (BWA)[7] v0.7.17. Duplicates were removed using Picard v2.18.17 (http://broadinstitute.github.io/picard/) and base quality was adjusted with GATK v 4.1.6. SNVs and indels were called using HaplotypeCaller in GATK and variant quality control filtering was applied as per Broad Institute standard best practices. Resulting variants were annotated using SnpEff[8] (v4.3t) and ANNOVAR[9] (2019Oct24).

Germline variants in 166 cancer predisposition genes present on all sequencing platforms were further analyzed. Variants with read-depth coverage ≥ 15, variant allele fraction ≥ 0.20, and observed in < 0.1% across each population in the public control databases non-TCGA ExAC (exonic) or gnomAD v2.1 (non-exonic, splicing) were included in the study.

##### *Assessment of variant pathogenicity*

Pathogenicity of retained variants was assessed *in silico* using custom software to evaluate ClinVar (11-25-2020) and a modified execution of InterVar (11-25-2020) (eFigure 1). First, ClinVar calls were considered in a hierarchical manner: (1) expert panel decision, (2) consensus of "badged labs", when available. Badged labs were defined as clinical laboratories meeting minimum requirements for data sharing to support quality assurance, as defined by ClinGen (https://www.clinicalgenome.org/tools/clinical-lab-data-sharing-list/ downloaded 12-2020). Next, prior to running InterVar, we adjusted PP5 based on this modified ClinVar assessment and corrected PVS1 using AutoPVS1[10]. Variants were then assigned to be pathogenic (P), likely pathogenic (LP), benign (B), likely benign (LB) or variants of unknown significance (VUS) by first considering the adjusted ClinVar results and then the modified InterVar output. This approach was applied to both neuroblastoma cases and controls  without cancer samples (PMBB and gnomAD 2.1).

##### *Sanger sequencing*

To verify germline variants, primers were designed with Primer3 and PCR reactions were carried out on 25 ng of DNA using optimized conditions for each reaction. Products were checked via gel electrophoresis. If the product had multiple bands, the entire remaining sample was run out then bands of interest excised and the DNA extracted using MinElute Gel Extraction Kit from Qiagen. Products with single bands were cleaned up and prepared for sequencing using the MinElute PCR Purification Kit (Qiagen). Samples were sequenced with two picomoles of the same primer used to create the amplicon.

##### *Germline copy number variant (CNV) analyses*

Rare germline CNVs affecting *BARD1* were identified using a SNP genotyping dataset of 5,585 neuroblastoma patients and 23,505 cancer-free control children as previously described[11]. Briefly, patients diagnosed with neuroblastoma or ganglioneuroblastoma were recruited through the Children's Oncology Group (COG) ANBL00B1 biology study without selection for clinical presentation. Germline DNA was isolated from peripheral blood or bone marrow mononuclear cells at time of diagnosis. Control children were recruited through the Children's Hospital of Philadelphia (CHOP) and screened for cancer and severe neurological or immunological disorders. Cases and controls were genotyped at the CHOP Center for Applied Genomics on matched Illumina SNP arrays and filtered for cryptic relatedness. CNVs were called using Nexus Copy Number 8.0 with linear correction for GC content, requiring a minimum of 10 probes per CNV. B-Allele Frequency (BAF) and Log R Ratio (LRR) plots were visually inspected to rule out potential artifacts.

##### *Somatic variant analyses*

Subjects harboring a germline P-LP variant in a cancer-predisposition gene were further assessed using matched tumor sequencing data when available. Specifically, somatic variants previously reported by Brady and colleagues[12] and additional variants from CAP sequencing data generated through TARGET were interrogated to assess potential second hits involving somatic SNVs, translocations, and focal copy number variations. For the TARGET CAP data, 499 tumor-normal pairs were analyzed. Somatic SNV calling was performed using Mutect 1.1.4 and annotated using Variant Effect Predictor (VEP). Somatic copy number variant (CNV) calling was performed using an in-house tool called VisCap, (https://github.com/pughlab/VisCapCancer). Tumor and normal bam files were first run through depth of coverage tool by GATK v3.0.0 and depth of coverage output was used as input for the VisCap somatic copy number calling. The calls for CNV were categorized into focal, subgenic and broad. A focal copy number variation is a case where the whole gene has gone through a copy number change. A subgenic change is when a few exons of the genes has copy number change and a broad change is for multiple genes undergoing copy number change. As not all of CPGs were included in somatic CAP sequencing for TARGET, we evaluated tumor DNA sequencing from a total of 79 subjects with germline P-LP variant in CPGs (20 cases with capture sequencing and all (n=59) cases with WES and/or WGS tumor data) were assessed for single nucleotide variation. However, all cases with germline P-LP variants in CPGs were included in tumor DNA copy number analyses.

***Statistical analyses***

The prevalence of P-LP germline variants in cancer predisposition genes was reported for the neuroblastoma cohort as a whole and within clinical and biological subsets. Fisher's exact test was used to compare clinical characteristics of patients with and without P-LP germline variants in the genes studied. A two-sided Fisher's exact test ($p$-value $< 0.05$) was considered significant. Enrichment testing of P-LP variants in cases vs. controls was performed at the overall, gene, and pathway level using Fisher's exact and corresponding odds ratios (ORs) and 95% confidence intervals were computed. Where zeros cause problems with computation of the odds ratio or its confidence intervals, the Woolf logit approach was utilized. A Bonferroni correction was applied to account for multiple testing at the gene level. Kaplan-Meier analyses of event-free and overall survival were performed to compare outcomes of patients with and without germline P-LP variants. A log-rank $p$-value $< 0.05$ was considered to be statistically significant. Statistical analyses were performed using R, version 3.3.2 with R Studio, version 1.0.136. Multivariate analyses were performed with a Cox proportional-hazards regression model to identify variables that were independently predictive of outcome. A $p$-value $< 0.05$ was considered to indicate statistical significance. Variables considered included: presence of germline P-LP variant, age at diagnosis, INSS stage, *MYCN* amplification status, and risk group. The patient cohort analyzed (n=774) included all those for whom complete data were available for the variables in the model.

#### II. SUPPLEMENTARY TABLES

##### eTable 1. Neuroblastoma patient clinical, tumor biologic and sequencing data (Excel file)

This gene panel was designed to capture known cancer predisposition genes (CPGs), genes implicated in neuroblastoma-associated syndromes (syndrome), genes identified through the neuroblastoma genome-wide association study (WGS). CPG: Cancer Predisposition Gene. Chromosome location: hg19.

##### eTable 2. Cancer predisposition genes panel (n=166) used in study (Excel File)

This gene panel was designed to capture known cancer predisposition genes (CPGs), genes implicated in neuroblastoma-associated syndromes (syndrome), genes identified through the neuroblastoma genome-wide association study (WGS). CPG: Cancer Predisposition Gene. Chromosome location: hg19.

##### eTable 3. Cancer predisposition gene P-LP variants observed in neuroblastoma (Excel File)

#### eTable 4. Neuroblastoma cases harboring multiple germline P-LP variants in CPGs

| Subject Identifier | COG Risk Group | *MYCN* status | Sex | Gene | Variant | Pathogenicity |
|---|---|---|---|---|---|---|
| PAPRXW | High | Amplified | Male | *ERCC2* | NM_000400:c.C2150G:p.A717G | P: ClinVar |
| | | | | *FANCA* | NM_001286167:c.189+1G>T | P: InterVar |
| | | | | *GJB2* | NM_004004:c.G416A:p.S139N | P: ClinVar |
| PARSEA | High | Not Amplified | Male | *BARD1* | NM_000465:c.C448T:p.R150X | P: ClinVar |
| | | | | *ERCC5* | NM_000123:c.2604_2605del:p.T868fs | LP: InterVar |
| PASNEF | High | Not Amplified | Female | *BRCA1* | NM_007297:c.G5101T:p.G1701C | LP: ClinVar |
| | | | | *BRCA2* | NM_000059:c.G3922T:p.E1308X | P: ClinVar |
| PATPXJ | High | Not Amplified | Female | *DOCK8* | NM_001190458:c.3654delC:p.D1218fs | LP: InterVar |
| | | | | *GBA* | NM_001005741: c.-50G>C | P: InterVar |
| PATVTL | High | Amplified | Male | *BRCA2* | NM_000059:c.5067dupA:p.A1689fs | P: ClinVar |
| | | | | *MSH3* | NM_002439:c.2759delC:p.S920fs | P: ClinVar |
| PAUICI | High | Not Amplified | Female | *IKZF1* | NM_001291845:c.T479A:p.L160X | LP: InterVar |
| | | | | *PHOX2B* | NM_003924:exon3:c.430-2A>G | P: InterVar |

#### eTable 5. Primers for Sanger sequencing validation germline variants

| Gene | Subject | Location (hg19) | Type | Ref | Alt | Forward Primer | Reverse Primer |
|---|---|---|---|---|---|---|---|
| *ATM* | PARNCW | 11:108165753 | missense | G | T | CATTGTAGGGTTTGCAGTGGA | TGGCAGAGGATGAATAAAACAGG |
| *BARD1* | PATGWT | 2:215595215 | stop gain | C | T | TCCTGGCTTAGGTTTTTCAGA | GCAATGTTCAAGATGCCAAA |
| *BARD1* | PASFDU | 2:215610566 | stop gain | G | T | GATGCCCTGGGTATAGAGAGC | TCTACCCCACCTCCCAAAATTC |
| *BARD1* | PATHJZ | 2:215645738 | frameshift | T | --- | GAGGGCTAAAAAGGCTTCTGC | TTTCTGAGGGCACCGTTTGC |
| *BARD1* | PARSEA | 2:215646150 | stop gain | C | T | AAATTCTTCGGGAGCTCCATGTG | TCAGAAACATCTGCAGGAGGAC |
| *BARD1* | PAHYWC | 2:215657051 | stop gain | C | T | AAGTGACTGCATTGGAACTGG | ATTCCAGAACTCCAGATAGATGTTT |
| *BRCA2* | PAPZYP | 13:32937375 | missense | A | G | ACACTGCTGTTCTCCTGTCA | ACAGCATACCACCCATCTGT |
| *CHEK2* | PARNEE | 22:29121242 | missense | G | A | CGCCCAGCAACTTACTCATC | GCCCTCTGATGCATGCTTTT |
| *ERCC2* | PAPZYP | 19:45868346 | missense | G | A | GGGCATCAAATTCCTGGGAC | AAGTTGTCCAAAACCCCAGC |
| *EZH2* | PASEGA | 7:148511124 | deletion | CAAGT | C | GGGTGCATTACCCAGAGAAA | AGGTGGTTGTGAGGGTTGAG |
| *EZH2* | PASTXV | 7:148515025 | deletion | C | --- | AAATCCAATCGGCAAAACAC | AGAACTTTGCCCTGATGTTGA |
| *EZH2* | PAVCJZ | 7:148544391 | deletion | ATG | --- | ATTTAGGGAGGCATTTCTGC | TGGCCGCAATTTAGTGTAGA |
| *EZH2* (Somatic) | PASEGA | 7:148525988-7:148531881 | deletion | 5893bp | --- | CCACCCTACCTGGCCATAAT | ATTAAGCTCACGGGTGTTGC |
| *FANCD2* | PASJYB | 3:10076378 | splice | G | C | ACACCCTTCCTATCCCAAAGT | TGAAACAACTGTGCTCTCCC |
| *WRN* | PARACS | 8:30924657 | missense | C | G | GAAGGCTATCTGTGGGTTGTATT | AGCCTGGATTTATTAGCCTTTCA |

**eTable 6. Gene-based testing for excess burden of P-LP variants vs. PMBB (Excel File)**
**eTable 7. Gene-based testing for excess burden of P-LP variants vs. gnomAD 2.1 (Excel File)**

**eTable 8. P-LP variants in genes with excess burden in neuroblastoma.**

| Gene[a] | Subject Identifier | Gender | COG Risk Group | *MYCN* status | Variant[b] | AA Change | Variant Type |
|---|---|---|---|---|---|---|---|
| *ALK* | PANYGR | Male | High | Not Amplified | c.3749T>C | I1250T | Missense |
| *ALK* | PARVLK | Female | High | Not Amplified | c.3824G>A | R1275Q | Missense |
| *ALK* | PATDVF | Female | Intermediate | Not Amplified | c.3824G>A | R1275Q | Missense |
| *BARD1* | PARSEA | Male | High | Not Amplified | c.448C>T | R150X | Stopgain |
| *BARD1* | PATGWT | Male | High | Amplified | c.1921C>T | R641X | Stopgain |
| *BARD1* | PAHYWC | Male | High | Amplified | c.334C>T | R112X | Stopgain |
| *BARD1* | PATHJZ | Female | Intermediate | Not Amplified | c.860_861del | E287fs | Frameshift |
| *BARD1* | PASFDU | Female | High | Not Amplified | c.1690C>T | Q564X | Stopgain |
| *BARD1* | PASGEE | Male | High | Not Amplified | c.1677+1G>T | - | Splice |
| *BARD1* | PATZRU | Male | High | Not Amplified | c.159-1G>T | - | Splice |
| *BARD1* | PASCIX | Male | High | Not Amplified | c.1954_1955insTGAACAGGAA GAAAAGTATG | E652fs | Frameshift |
| *EZH2* | PASEGA | Male | High | Not Amplified | c.1774_1777delACTT | T592fs | Frameshift |
| *EZH2* | PASTXV | Male | Low | Not Amplified | c.1184delG | G395fs | Frameshift |
| *EZH2* | PAPVXS | Male | High | Not Amplified | c.625G>A | D209N | Missense |
| *EZH2* | PAVCJZ | Male | High | Not Amplified | c.-1_2delCAT | M1del | Start Lost |

#### eTable 9. Rare germline copy number variants (CNVs) disrupting *BARD1* in neuroblastoma cases

##### *SNP array CNV calls*

| Sample | CN Change | Estimated Size | Estimated Region | Min Length | Min Region (hg19) | Max Length | Max Region | Probe Median | Probe Count |
|---|---|---|---|---|---|---|---|---|---|
| PAVNLD | CN Loss | 168822 | chr2:215487324-215656145 | 161451 | chr2:215489130-215650580 | 176192 | chr2:215485519-215661710 | -0.39509578 | 36 |
| PAWNMH | CN Loss | 182685 | chr2:215549453-215732137 | 153593 | chr2:215562577-215716169 | 211777 | chr2:215536329-215748105 | -0.436415702 | 39 |
| PALXTB | CN Loss | 70165 | chr2:215623659-215693823 | 49912 | chr2:215627397-215677308 | 90418 | chr2:215619921-215710338 | -0.707099229 | 16 |

##### *Validation: WGS (Complete Genomics) Structural Variant Calls*

| Sample | SV Type | Size | Region (hg19) | Strand | Mate Pair Count | Frequency In Baseline Genome Set |
|---|---|---|---|---|---|---|
| PALXTB | Deletion | 61875 | chr2:215622570-215684445 | + | 20 | 0 |

##### *Validation: WGS (Complete Genomics) Copy Number Variation Calls*

| Sample | CNV Type | Size | Region (hg19) | Average Coverage | Relative Coverage | CNV Score (Phred-Scaled) |
|---|---|---|---|---|---|---|
| PALXTB | CN Loss | 62000 | chr2:215622000-215684000 | 25 | 0.5 | 53 |

#### eTable 10. Association of P-LP variants with clinical and tumor characteristics (Full Cohort)

| Characteristic | Without P-LP Variants (n=677) # subjects | Without P-LP Variants (n=677) % subjects | With P-LP variants (n=109) # subjects | With P-LP variants (n=109) % subjects | P-value |
|---|---|---|---|---|---|
| Sex | | | | | |
| Male | 384 | 57% | 63 | 58% | 0.9171 |
| Female | 293 | 43% | 46 | 42% | |
| Age at Diagnosis | | | | | |
| < 18 months | 206 | 30% | 36 | 33% | 0.5780 |
| > 18 months | 471 | 70% | 73 | 67% | |
| COG Risk Group | | | | | |
| Low | 91 | 13% | 12 | 11% | |
| Intermediate | 100 | 15% | 19 | 17% | 0.6494 |
| High | 486 | 72% | 78 | 72% | |
| INRG Stage | | | | | |
| Stage L1 | 77 | 16% | 8 | 12% | |
| Stage L2 | 72 | 15% | 10 | 15% | 0.6157 |
| Stage MS | 39 | 8% | 3 | 5% | |
| Stage M | 295 | 61% | 44 | 68% | |
| INSS Stage | | | | | |
| Stage 1 | 34 | 5% | 4 | 4% | |
| Stage 2a | 21 | 3% | 3 | 3% | |
| Stage 2b | 35 | 5% | 1 | 1% | 0.2187 |
| Stage3 | 83 | 12% | 10 | 9% | |
| Stage 4 | 460 | 68% | 86 | 79% | |
| Stage 4s | 44 | 6% | 5 | 5% | |
| MYCN status | | | | | |
| Amplified | 201 | 30% | 23 | 21% | 0.0672 |
| Not amplified | 467 | 70% | 85 | 79% | |
| Histologic Classification | | | | | |
| Favorable | 180 | 27% | 31 | 28% | 0.5491 |
| Unfavorable | 455 | 67% | 67 | 61% | |
| unknown | 42 | 6% | 11 | 10% | |
| Degree of Differentiation | | | | | |
| Differentiated | 30 | 5% | 5 | 5% | 0.8023 |
| Undifferentiated or poorly differentiated | 572 | 95% | 89 | 95% | |
| Mitosis Karyorrhexis Index | | | | | |
| Low | 228 | 39% | 35 | 39% | |
| Intermediate | 167 | 29% | 33 | 37% | 0.2276 |
| High | 184 | 32% | 22 | 24% | |
| Ploidy | | | | | |
| Diploid | 251 | 38% | 42 | 39% | 0.8307 |
| Hyperdiploid | 408 | 62% | 65 | 61% | |
| 1p LOH | | | | | |
| Yes | 193 | 34% | 27 | 33% | >0.9999 |
| No | 377 | 66% | 54 | 67% | |
| 11q LOH | | | | | |
| Yes | 129 | 23% | 29 | 36% | 0.0119* |
| No | 438 | 77% | 51 | 64% | |

#### eTable 11. Association of P-LP variants with clinical and tumor characteristics (Capture Only)

| Characteristic | Without P-LP variants (n=434) # subjects | Without P-LP variants (n=434) % subjects | With P-LP variants (n=55) # subjects | With P-LP variants (n=55) % subjects | P-value |
|---|---|---|---|---|---|
| Sex | | | | | |
| Male | 234 | 54% | 33 | 60% | 0.4727 |
| Female | 200 | 46% | 22 | 40% | |
| Age at Diagnosis | | | | | |
| < 18 months | 180 | 41% | 30 | 55% | 0.0822 |
| > 18months | 254 | 59% | 25 | 45% | |
| COG Risk Group | | | | | |
| Low | 80 | 18% | 9 | 16% | |
| Intermediate | 87 | 20% | 18 | 33% | 0.0962 |
| High | 267 | 62% | 28 | 51% | |
| INRG Stage | | | | | |
| Stage L1 | 76 | 20% | 8 | 16% | |
| Stage L2 | 70 | 18% | 10 | 20% | 0.4961 |
| Stage MS | 26 | 7% | 1 | 2% | |
| Stage M | 212 | 55% | 31 | 62% | |
| INSS Stage | | | | | |
| Stage 1 | 34 | 8% | 4 | 7% | |
| Stage 2a | 21 | 5% | 3 | 5% | |
| Stage 2b | 34 | 8% | 1 | 2% | 0.4352 |
| Stage 3 | 76 | 18% | 10 | 18% | |
| Stage 4 | 243 | 56% | 36 | 65% | |
| Stage 4s | 26 | 6% | 1 | 2% | |
| MYCN status | | | | | |
| Amplified | 128 | 30% | 9 | 17% | 0.0536 |
| Not Amplified | 303 | 70% | 45 | 83% | |
| Histologic Classification | | | | | |
| Favorable | 154 | 35% | 25 | 45% | 0.0954 |
| Unfavorable | 264 | 60% | 26 | 47% | |
| unknown | 16 | 4% | 4 | 7% | |
| Degree of Differentiation | | | | | |
| Differentiated | 19 | 5% | 3 | 6% | 0.7283 |
| Undifferentiated or poorly differentiated | 394 | 95% | 50 | 94% | |
| Mitosis Karyorrhexis Index | | | | | |
| Low | 164 | 41% | 21 | 41% | |
| Intermediate | 111 | 28% | 20 | 39% | 0.1239 |
| High | 126 | 31% | 10 | 20% | |
| Ploidy | | | | | |
| Diploid | 151 | 36% | 18 | 33% | 0.7645 |
| Hyperdiploid | 270 | 64% | 36 | 67% | |
| 1p LOH | | | | | |
| Yes | 142 | 33% | 19 | 35% | 0.7624 |
| No | 284 | 67% | 35 | 65% | |
| 11q LOH | | | | | |
| Yes | 79 | 19% | 17 | 31% | 0.0309* |
| No | 346 | 81% | 37 | 69% | |

#### eTable 12. Cox proportional hazards regression model results.

| Variable[1] | Coefficient (95% Confidence Interval) | P-value | Hazard Ratio (95% Confidence Interval) |
|---|---|---|---|
| P-LP Variant | 0.3677 (0.0648-0.6706) | 0.0174 | 1.4444 (1.0699-1.9554) |
| Age at Diagnosis | 0.0000 (-0.0001-1.0001) | 0.9251 | 1.0000 (0.9999-1.0001) |
| Stage 4 | 1.3102 (0.7826-1.8378) | < 0.0001 | 3.7069 (2.1871-6.2825) |
| MYCN amplification | 0.2610 (0.0021-0.5199) | 0.0481 | 1.2982 (1.0021-1.6818) |
| Risk Group | 1.0101 (0.5363-1.4839) | < 0.0001 | 2.7459 (1.7097-4.4100) |

[1] Model based on neuroblastoma patients with data on all variables (n=774).

#### III. SUPPLEMENTARY FIGURES

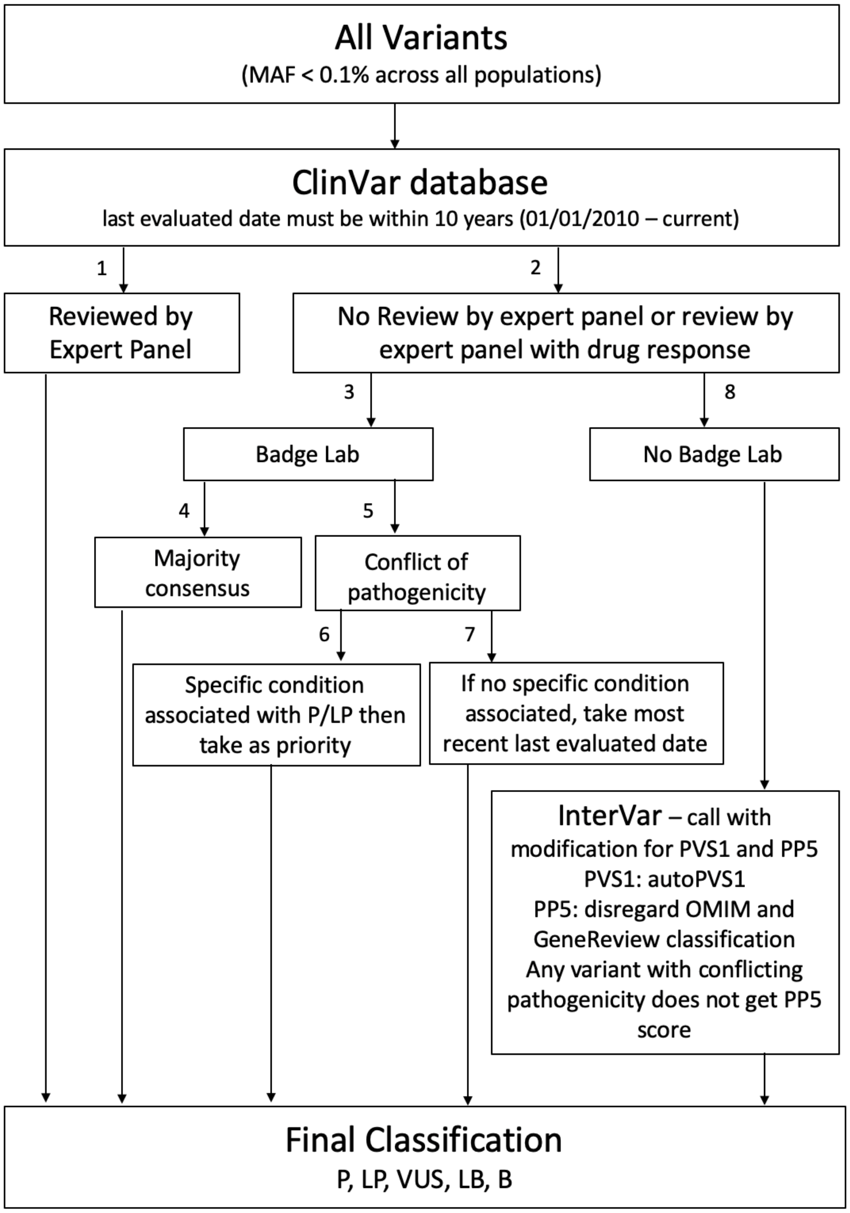

**eFigure 1**. **Flow diagram of automated pathogenicity assessment for germline variants.** Order of flow indicated by labeled arrows.

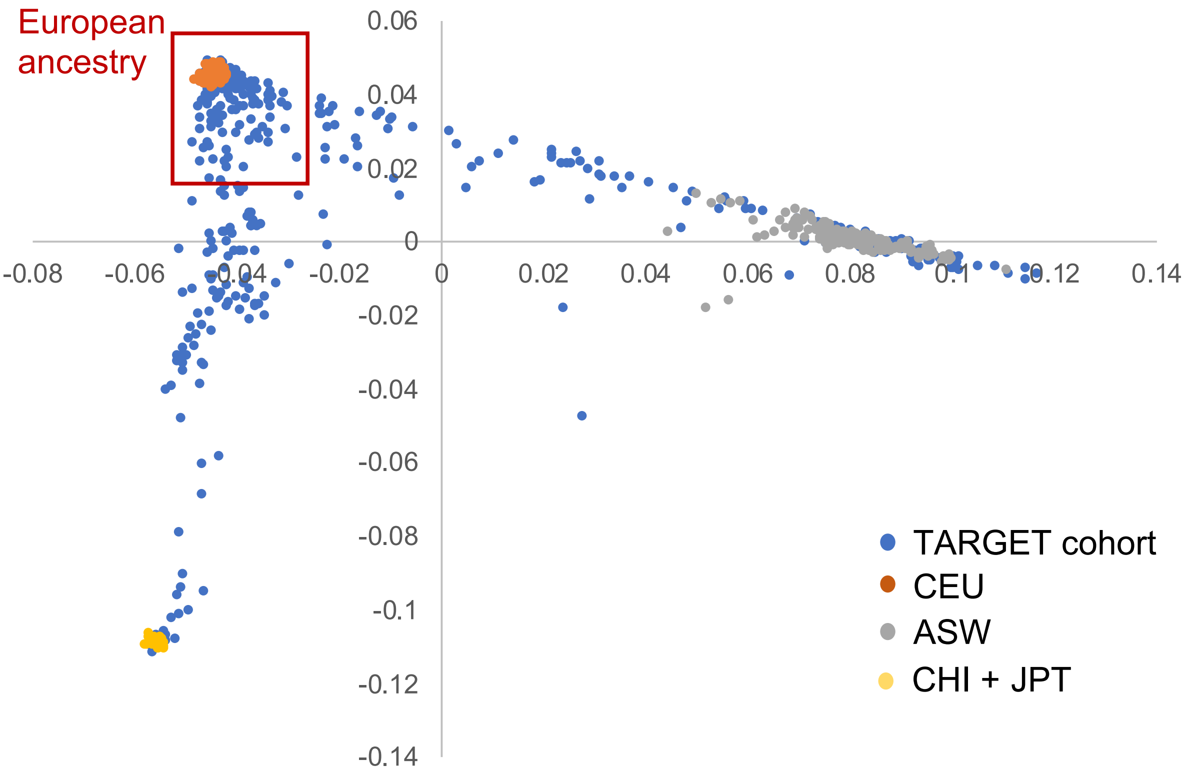

**eFigure 2. Ancestry of neuroblastoma cohort inferred from principal component analysis.**
Matched germline Illumina SNP array data were utilized to infer ancestry using principal component analysis of pruned SNPs together with the data from 1000 Genomes.

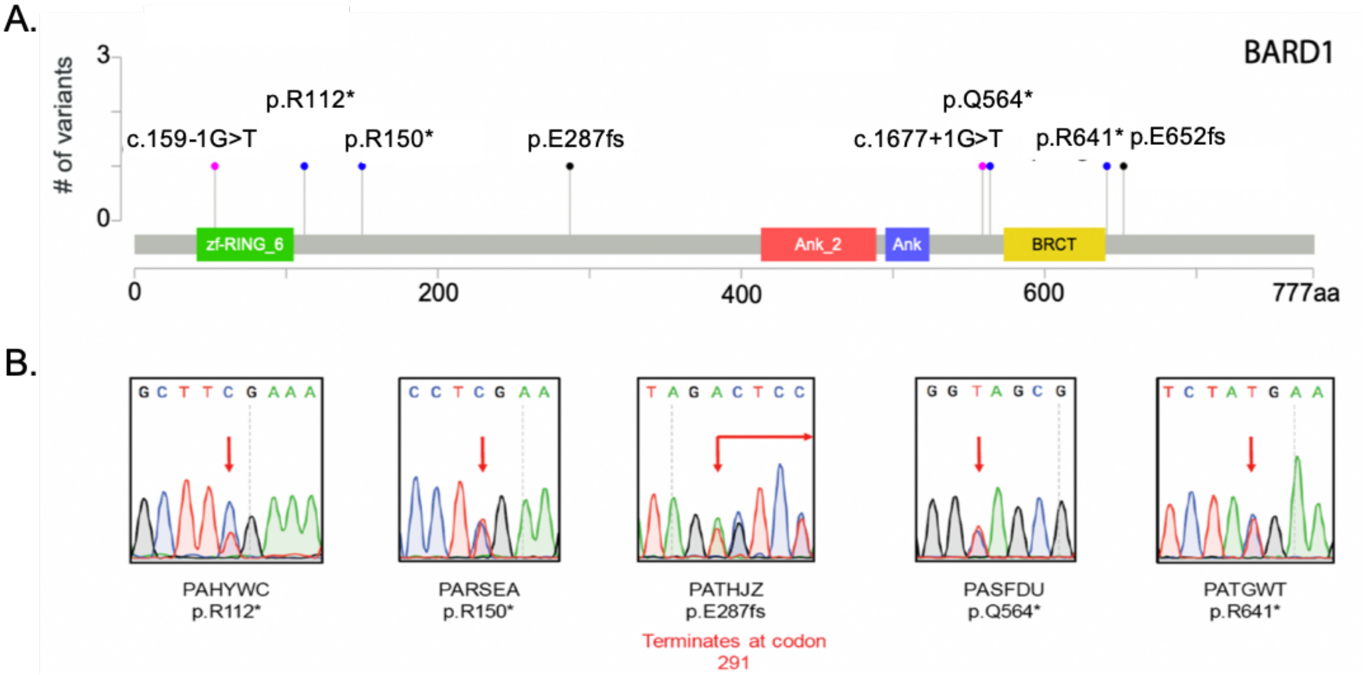

**eFigure 3. *BARD1* germline variants validated by Sanger sequencing. A.** Lollipop figure depicting pathogenic germline variants in neuroblastoma cases. **B.** Sanger validation of pathogenic germline variants in neuroblastoma. See **eTable 5** for primers used for Sanger sequencing. Variant annotations with respect to ENST00000260947.

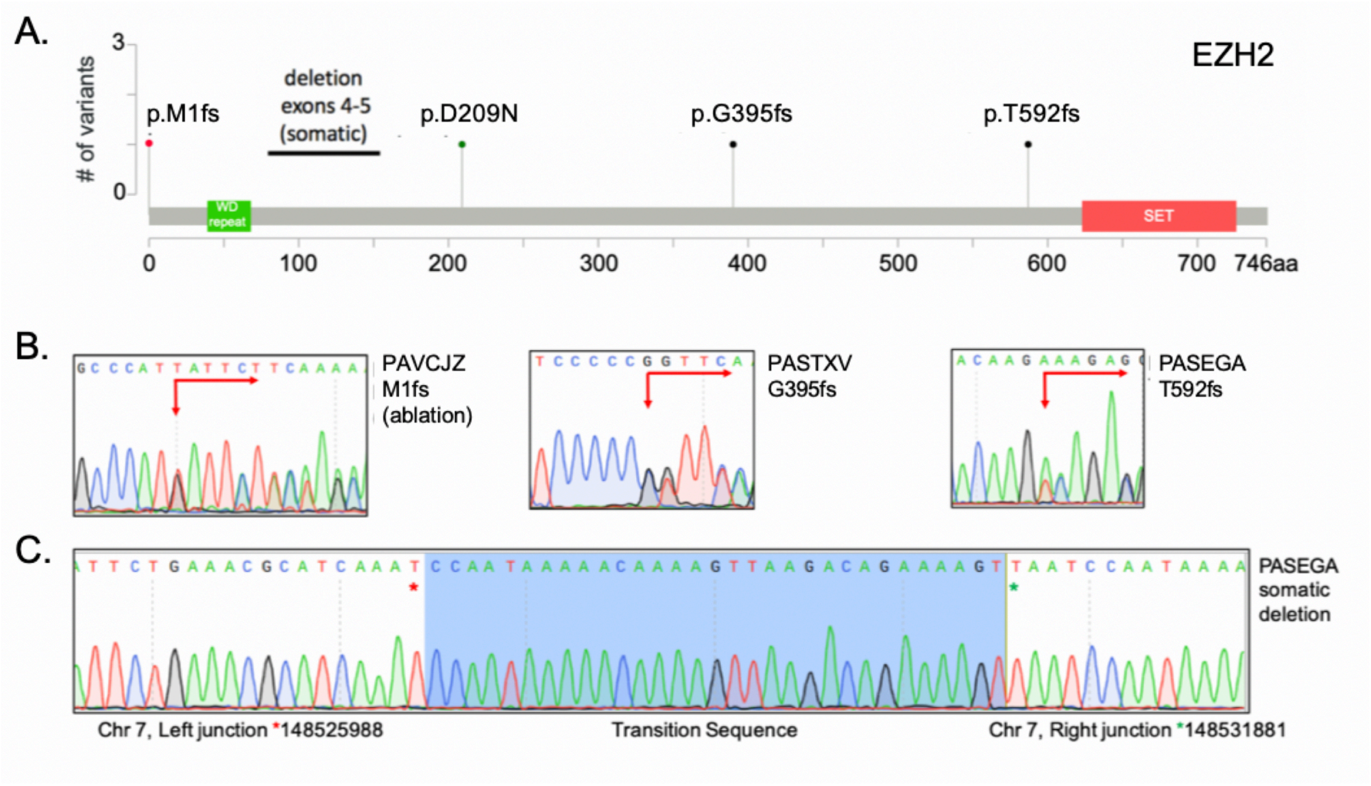

**eFigure 4. *EZH2* P/LP germline and somatic variants validated by Sanger sequencing. A.** Lollipop figure depicting pathogenic germline and somatic variants in neuroblastoma cases. **B.** Sanger validation of pathogenic germline and somatic variants in neuroblastoma patients PAVCJZ, PASTXV, and PASEGA. **C.** Sanger validation of *EZH2* somatic deletion in PASEGA. See **eTable 5** for primers used. Variant annotations with respect to ENST00000320356.

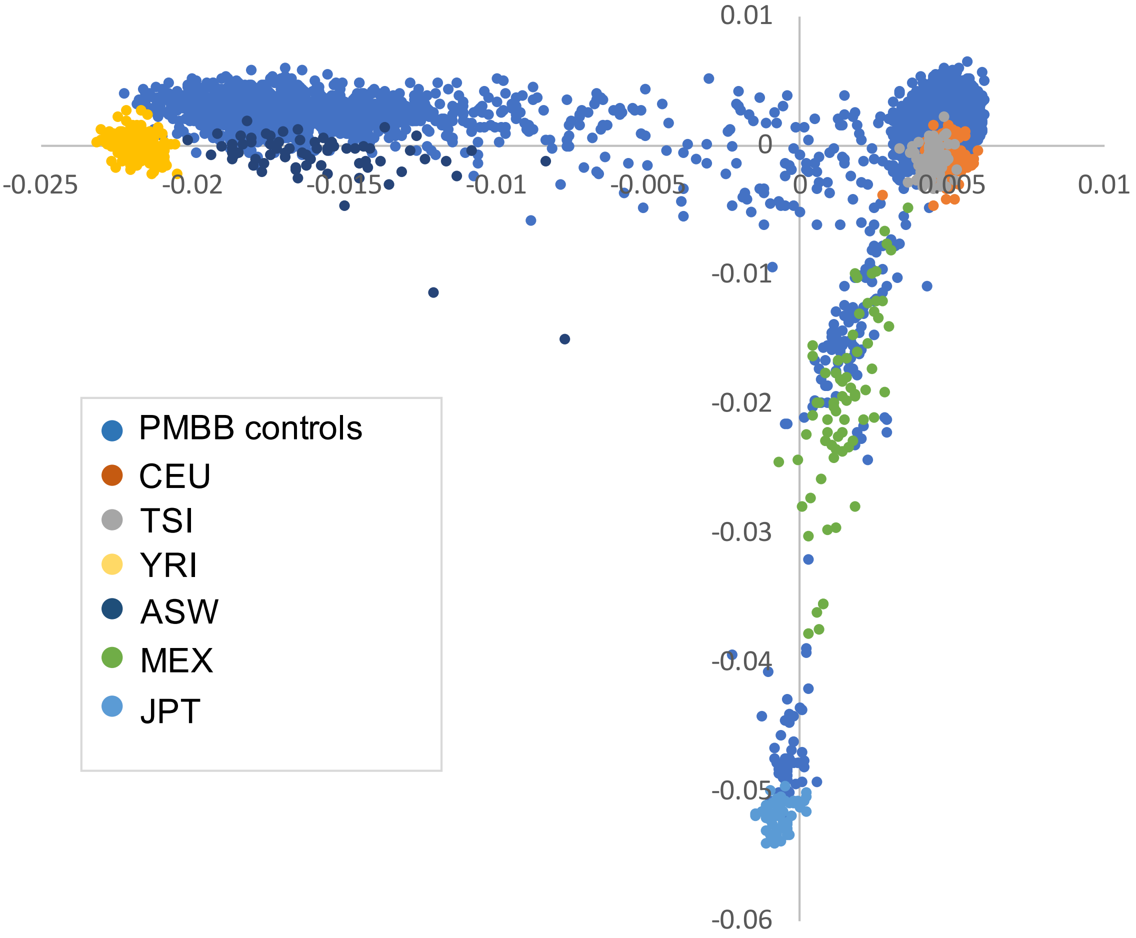

**eFigure 5. Ancestry of PMBB cohort inferred from principal component analysis.**

### IV. BANNER AUTHOR LISTS AND CONTRIBUTIONS

#### Penn Medicine BioBank Banner Author List and Contribution Statements

**PMBB Leadership Team**
Daniel J. Rader, M.D., Marylyn D. Ritchie, Ph.D., Michael D. Feldman M.D.

Contribution: All authors contributed to securing funding, study design and oversight. All authors reviewed the final version of the manuscript.

**Patient Recruitment and Regulatory Oversight**
JoEllen Weaver, Afiya Poindexter, Ashlei Brock, Khadijah Hu-Sain, Yi-An Ko

Contributions: JW manage patient recruitment and regulatory oversight of study. AP, AB, KH, YK recruitment and enrollment of study participants.

**Lab Operations**
JoEllen Weaver, Meghan Livingstone, Fred Vadivieso, Ashley Kloter, Stephanie DerOhannessian, Teo Tran, Linda Morrel, Ned Haubein, Joseph Dunn

Contribution: JW, ML, FV, SD oversight of lab operations. ML, FV, AK, SD, TT, LM perform sample processing. NH, JD are responsible for sample tracking and the laboratory information management system.

**Clinical Informatics**
Anurag Verma, Ph.D., Colleen Morse, M.S., Marjorie Risman, M.S., Renae Judy, B.S.

Contribution: All authors contributed to the development and validation of clinical phenotypes used to identify study subjects and (when applicable) controls.

**Genome Informatics**
Anurag Verma Ph.D., Shefali S. Verma, Ph.D., Yuki Bradford, M.S., Scott Dudek, M.S., Theodore Drivas, M.D., PH.D.

Contribution: A.V., S.S.V. are responsible for the analysis design and infrastructure needed quality control genotype and exome data. Y.B. performed the analysis. T.D. and A.V. provide variant and gene annotations and their functional interpretation of variants.

#### Regeneron Genetics Center Banner Author List and Contribution Statements

**RGC Management and Leadership Team**
Goncalo Abecasis, Aris Baras, Michael Cantor, Giovanni Coppola, Andrew Deubler, Aris Economides, Katia Karalis, Luca A. Lotta, John D. Overton, Jeffrey G. Reid, Katherine Siminovitch & Alan Shuldiner

**Sequencing and Lab Operations**
Christina Beechert, Caitlin Forsythe, Erin D. Fuller, Zhenhua Gu, Michael Lattari, Alexander Lopez, John D. Overton, Maria Sotiopoulos Padilla, Manasi Pradhan, Kia Manoochehri, Thomas D. Schleicher, Louis Widom, Sarah E. Wolf & Ricardo H. Ulloa

**Clinical Informatics**
Amelia Averitt, Nilanjana Banerjee, Michael Cantor, Dadong Li, Sameer Malhotra, Deepika Sharma & Jeffrey C. Staples

**Genome Informatics**
Xiaodong Bai, Suganthi Balasubramanian, Suying Bao, Boris Boutkov, Siying Chen, Gisu Eom, Lukas Habegger, Alicia Hawes, Shareef Khalid, Olga Krasheninina, Rouel Lanche, Adam J. Mansfield, Evan K. Maxwell, George Mitra, Mona Nafde, Sean O'Keeffe, Max Orelus, Razvan Panea, Tommy Polanco, Ayesha Rasool, Jeffrey G. Reid, William Salerno, Jeffrey C. Staples, Kathie Sun & Jiwen Xin

**Analytical Genomics and Data Science**
Goncalo Abecasis, Joshua Backman, Amy Damas, Lee Dobbyn, Manuel Allen Revez Ferreira, Arkopravo Ghosh, Christopher Gillies, Lauren Gurski, Eric Jorgenson, Hyun Min Kang, Michael Kessler, Jack Kosmicki, Alexander Li, Nan Lin, Daren Liu, Adam Locke, Jonathan Marchini, Anthony Marcketta, Joelle Mbatchou, Arden Moscati, Charles Paulding, Carlo Sidore, Eli Stahl, Kyoko Watanabe, Bin Ye, Blair Zhang & Andrey Ziyatdinov

**Research Program Management & Strategic Initiatives**
Marcus B. Jones, Jason Mighty & Lyndon J. Mitnaul
